# The variable genomic landscape during osteosarcoma progression: insights from a longitudinal WGS analysis

**DOI:** 10.1101/2024.04.18.24306025

**Authors:** Debora M. Meijer, Dina Ruano, Inge H. Briaire-de Bruijn, Pauline M. Wijers-Koster, Michiel A.J. van de Sande, Hans Gelderblom, Anne-Marie Cleton-Jansen, Noel F.C.C. de Miranda, Marieke L. Kuijjer, Judith V.M.G. Bovée

**Author notes:** These authors contributed equally to this work (Marieke L. Kuijjer, Judith V.M.G. Bovée).

## Abstract

Osteosarcoma is a primary bone tumor that exhibits a complex genome characterized by gross chromosomal abnormalities. Osteosarcoma patients often develop metastatic disease, resulting in limited therapeutic options and poor survival rates. To gain knowledge on the mechanisms underlying osteosarcoma heterogeneity and metastatic process, it is important to obtain a detailed profile of the genomic alterations that accompany osteosarcoma progression. We performed WGS on multiple tissue samples from six patients with osteosarcoma, including the treatment naïve biopsy of the primary tumor, resection of the primary tumor after neoadjuvant chemotherapy, local recurrence and distant metastases. SNVs and structural variants were found to accumulate over time, contributing to an increased complexity of the genome of osteosarcoma during progression. Phylogenetic trees based on SNVs and structural variants reveal distinct evolutionary patterns between patients, including linear, neutral and branched patterns. The majority of osteosarcomas showed variable copy number profiles or gained whole genome doubling in later occurrences. Additionally, chromothripsis is not confined to a single early event, as multiple other chromothripsis events may appear in later occurrences. Together, we provide a detailed analysis of the complex genome of osteosarcomas and show that five out of six osteosarcoma genomes are highly dynamic and variable during progression.

## Introduction

Osteosarcoma is the most common primary malignant bone tumor, occurring mostly in children and young adults [1]. Current treatment options are limited to (neo)adjuvant conventional chemotherapy and surgical resection. Osteosarcomas show high genetic heterogeneity. They exhibit a complex genomic landscape characterized by large chromosomal abnormalities, including copy number alterations (CNAs) and structural variants [2, 3]. The number of small variants, such as single nucleotide variants (SNVs), is relatively low compared to tumor types that often arise due to mutagen exposure, such as lung cancer and melanoma [4]. Recurrent alterations include mutations in *TP53* or *RB1*, occurring in >90% and 56% of sporadic osteosarcomas, respectively [1]. Osteosarcomas also frequently display chromothripsis, a phenomenon involving the acquisition of tens to hundreds of genomic rearrangements in a confined region within a single event [5]. A study analyzing chromothripsis events from whole genome sequencing (WGS) data in a wide variety of cancer types found that 29 out of 34 primary osteosarcomas are affected by chromothripsis [6]. In the majority of osteosarcomas that harbor chromothripsis, events seem to affect random locations. However, recurrent chromothripsis events have been observed in 5-16% of patients affecting chromosomes 5, 12 and 17, possibly resulting in the selection of driver genes [7]. Chromothripsis in osteosarcoma has not been investigated yet in a longitudinal study and it is unknown whether these events can accumulate during progression.

The high interpatient heterogeneity makes it difficult to understand the tumorigenesis and metastatic process of osteosarcoma. Few studies have performed longitudinal genomic analysis to identify early drivers of osteosarcoma. A WES analysis of osteosarcoma tumors developing during the course of disease has shown that early mutations are frequently found in genes associated with cell cycle G1 transition, such as *TP53, RB1* and *CDKN2A* [8]. Alterations acquired in later occurrences often involve, amongst others, *MYC* and *BRCA2* [8]. Given that osteosarcomas are often treated with chemotherapy, another study found that pathogenic alterations in *NF1* and *KIT* can potentially be acquired as a consequence of cisplatin treatment, impacting the subsequent progression [9]. To further dissect the clonal and intrapatient heterogeneity of osteosarcoma, a multi-region sequencing study with paired primary-metastasis osteosarcoma samples revealed high heterogeneity within single primary tumor samples and a dynamic mutational process during osteosarcoma progression [10]. This would suggest a constant source of clonal heterogeneity, which makes it difficult for the immune system or therapeutic approaches to eliminate the tumor. Unfortunately, the measurements of heterogeneity and identification of potential driver genes are mainly based on SNVs and CNAs. Large structural variants and chromothripsis events are less often identified due to their more challenging detection process and the requirement of WGS data, despite their high significance in osteosarcoma.

In this study we aimed to gain novel insights into the tumorigenesis and progression of osteosarcoma by doing an in-depth characterization of the osteosarcoma genome during the course of disease from six patients. WGS data was generated for a total of 24 samples which allowed us to profile the genomic complexity in detail. We show that the osteosarcoma genome can be highly variable during progression, particularly in terms of structural variants, CNA and chromothripsis events.

## Methods

### Sample information

Fresh-frozen osteosarcoma tissue samples were collected from six patients with advanced disease (P1-P6; M/F = 3/3; mean age at diagnosis = 20.3, range = 14-30), including the treatment naïve biopsy of the primary tumor (N=6)), resection of the primary tumor after neoadjuvant chemotherapy (N=4), local recurrences (N=4 in three patients), distant metastases (N=10 in five patients) and paired normal tissue for each patient (table 1). Tumor percentage of the samples was at least 70% as estimated by an experienced pathologist (JVMGB). Matched RNA-seq of the same patient samples was previously published (GSE237033). Approval of institutional review board (Medisch-Ethische Toetsingscommissie Leiden Den Haag Delft) was obtained for the use of these tissue samples from the LUMC bone and soft tissue tumor biobank (Leiden, the Netherlands) (B20.067). Written informed consent from patients was obtained. All specimens were pseudoanonymized and handled according to the ethical guidelines described in ‘Code for Proper Secondary Use of Human Tissue in The Netherlands’ of the Dutch Federation of Medical Scientific Societies.

**Table 1.** Patient clinical information.

| Patient | Sex | PRM<sup>1</sup> | Location | Histotype | Status<sup>1</sup> | Treatment | Chemotherapy response |
| --- | --- | --- | --- | --- | --- | --- | --- |
| P1 | F | Primary B<br>Primary R<br>Recurrence 1 | Humerus<br>Humerus<br>Humerus | Fibroblastic | NED | Chemotherapy | Poor |
| P2 | F | Primary B<br>Recurrence 1<br>Recurrence 2<br>Metastasis 1 | Radius<br>Soft Tissue<br>Soft Tissue<br>Lung | Teleangiectatic | NED | Chemotherapy | Good |
| P3 | M | Primary B<br>Metastasis 1<br>Metastasis 2 | Femur<br>Lung<br>Sella | Osteoblastic | DOD | Chemotherapy | Poor |
| P4 | M | Primary B<br>Primary R<br>Metastasis 1<br>Metastasis 2<br>Metastasis 3<br>Metastasis 4<br>Metastasis 5 | Humerus<br>Humerus<br>Lung<br>Lung<br>Lung<br>Pancreas<br>Mesocolon | Osteoblastic | AWD | Chemotherapy | Poor |
| P5 | F | Primary B<br>Primary R<br>Metastasis 1 | Fibula<br>Fibula<br>Sternum | Osteoblastic | DOD | Chemotherapy | Poor |
| P6 | M | Primary B<br>Primary R<br>Recurrence 1<br>Metastasis 1 | Iliac Bone<br>Iliac Bone<br>Soft tissue<br>Soft tissue | Chondroblastic | AWD | Chemotherapy | Poor |

<sup>1</sup> Patient outcome status: no evidence of disease (NED); Dead of disease (DOD); Alive with disease (AWD). Primary B, Primary Biopsy; Primary R, primary Resection.

### WGS data generation and data pre-processing

DNA was isolated from fresh-frozen tissue using the Wizard Genomic DNA purification kit (A1125, Promega, Madison, WI, USA), according to manufacturer’s instructions. WGS was performed by GenomeScan (Leiden, the Netherlands) on the NovaSeq6000 platform producing 152bp paired-end reads with 90x depth for tumor DNA and 30X for normal DNA. The raw data was processed using a reimplementation of the Hartwig Medical Foundation (HMF) pipeline (v1.01, https://github.com/biowdl/WGSinCancerDiagnostics/releases/tag/v1.0.1). The pipeline was run using default settings and aligned against the GRCh37 reference genome using the BWA algorithm.

### Variant calling

SNVs, SVs, CNAs and minor allele copy numbers were obtained using PURPLE [11], a computational tool that is part of the HMF pipeline. PURPLE estimates tumor ploidy and purity from a combination B-allele frequencies, read depth ratios, somatic variants and structural variants. Subsequently, PURPLE infers copy number profiles, recover low confidence structural variants, annotate somatic variants, adjust copy numbers for tumor purity and determine whole genome doubling (WGD) status. WGD was detected when at least 11 autosomes had a major allele copy number > 1.5 as described in the manual of PURPLE. Variants are reported by PURPLE if they are located on a driver gene, which is a panel of genes manually curated by Hartwig (DriverGenePanel.38.tsv, HMFtools-Resources) [11]. From the reported SNV and indels reported on l driver genes, only pathogenic variants (class 4 and class 5 according to ACMG variant classification guidelines) were selected for SNV quantification and subsequent analyses. Structural variant types were included as annotated by PURPLE: translocations, deletions, duplications, insertions, inversions, single breakend structural variants (SGL) and inferred structural variants to explain copy number discrepancies without suitable structural variant candidates. Potential driver genes per patient and the number of SNVs and structural variants were visualized in R (version 4.1.0) using R packages *pheatmap* and *ggplot2*. LOH regions were called when the minor allele copy number was < 0.1. Copy number and LOH profiles were visualized with R package *GenVisR* (version 1.22.1).

### Phylogenetic tree reconstruction

Phylogenetic trees were build using PAUP (version 4.0a169) [12]. For this, a binary matrix was made for each patient separately, containing the presence (1) or absence (0) of each SNV and SV per tumor. A nexus file was generated to specify parsimony parameters. The outgroup function was used to set an artificial normal sample, a sample containing only zeros in the binary matrix, as starting point. A heuristic search performed with default settings resulted in the consensus tree.

### Detection of chromothripsis events

Candidate chromothripsis events were identified using ShatterSeek according to developers’ criteria [6]. Amongst the criteria, chromothripsis events should contain at least 7 adjacent segments that oscillate between two copy number states. In order to detect candidate chromothripsis events lacking the typical oscillating copy number pattern, which is common in osteosarcomas, chromothripsis events were also called when at least 40 intraor interchromosomal structural variants were detected in a region, as described by Cortes-Ciriano et al. (2020). All candidate chromothripsis events were revised by visual inspection to refine the call set, as the complex genome of osteosarcomas could raise false positive events. Chromothripsis events were annotated as “canonical chromothripsis” when at least 60% of segments oscillated between two copy number states. All other events were annotated as “non-canonical chromothripsis”, lacking the typical oscillating copy number pattern. Additionally, missed potential chromothripsis events by ShatterSeek which were only detected by visual inspection were added to obtain the final call set. The majority of those missed events by ShatterSeek included events that almost met the criteria for chromothripsis and evidence for chromothripsis was detected in similar regions in other tumor samples of the same patient. Remaining missed regions that were added after visual inspection were characterized by multiple interchromosomal translocations involving chromosomes with chromothripsis called by ShatterSeek. Circos plots to visualize translocations and chromothripsis were made using the CIRCOS tool implemented in PURPLE.

### Gene fusion detection

STAR-Fusion (version 1.10.0) [13] and FusionCatcher (version 1.33) [14] were performed for gene fusion detection on matching RNA-seq data which is publicly available (GSE237033).

## Results

### Truncal and late driver alterations involved in osteosarcoma initiation and development

Detection of potential driver alterations during disease progression revealed genes that are likely involved in osteosarcoma initiation or development (supplementary Table S1). Truncal events included already known pathogenic alterations in genes such as *TP53, RB1, CDK4, MYC, SETD2* and *MDM2* (figure 1A). Shared truncal driver genes across the patients were detected in *TP53* (n = 3) and *SETD2* (n = 2). *TP53* was altered in different ways in three patients: a *TP53*::*CASK* gene fusion in P2, a *TP53* deletion (copy number = 0) in P3 and a *TP53* somatic missense mutation in P4. The gene fusion between *TP53* (exon 8) and *CASK* (exon 26) leads to a functional *TP53*::*CASK* fusion transcript, as confirmed using RNA-Seq data. *SETD2* was deleted in P5 and harbored a SNV in P6. Driver alterations in later occurrences, potentially providing a strong selective advantage, included SNVs in *NF1* and *PIK3CA*, and amplifications of *KDR, KIT* and *TERT*, although none of these altered genes were recurrent across patients.

**Figure 1.**
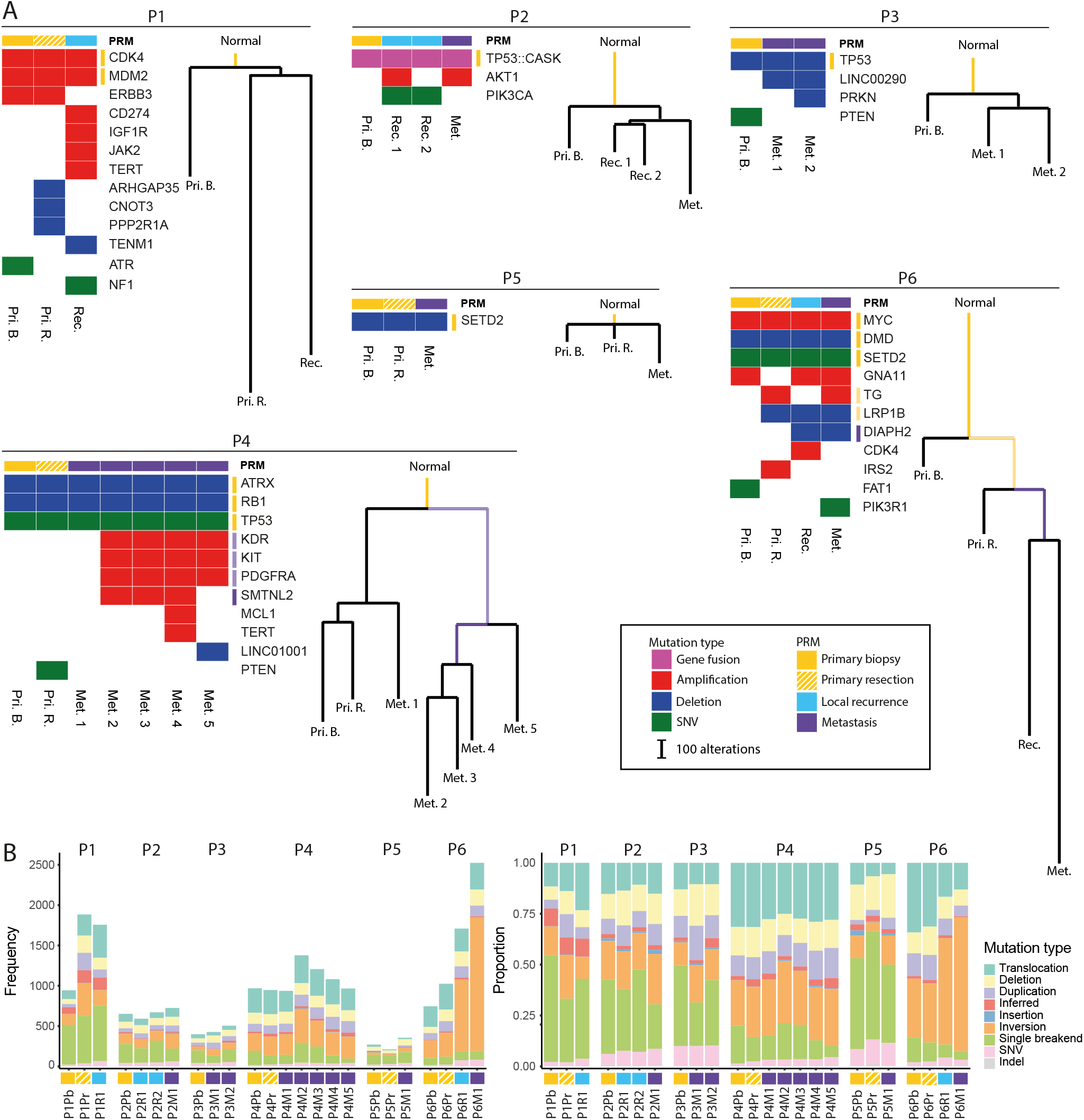
Driver genes, SNVs and SVs contribung to evoluonary paerns in osteosarcoma. (A) Phylogenec trees per paent generated by PAUP based on SNVs and structural variants visualizing evoluonary paerns. Length of the branches are proporonal to the number of mutaons. Driver genes are visualized in le plots and annotated based on amplificaons (red), deleons (blue) or SNVs (green). The acquirement of mutaons in driver genes during progression are indicated with yellow (truncal) and purple (late) to the specific locaon in the phylogenec tree. (B) The proporon and frequency of SNVs, indels and structural variant types for each tumor visualized in barplots. In the whole figure, the tumors of each paent are ordered from le to right based on date of diagnosis.

### Single nucleotide variants and structural variants reveal divergent evolutionary patterns

Phylogenetic trees based on SNVs and structural variants revealed variable evolutionary patterns between patients regarding the timing and selection of alterations (figure 1A). Following previously described models of tumor evolution [15], osteosarcoma tumor evolution showed patterns resembling linear, branched, neutral and (reversed) punctuated evolution. Interestingly, P1 showed relatively few truncal alterations and many unique alterations in later occurrences, displaying the opposite of a punctuated evolutionary pattern where a large number of truncal alterations may occur in a short period of time. P4 displayed a branched evolutionary pattern, where one branch with the primary biopsy, primary resection and first metastasis showed very distinct alterations compared to the other branch containing the second until fifth metastasis, on top of the truncal *ATRX, RB1* and *TP53* alterations. A linear evolutionary pattern was observed for P6, showing a stepwise acquisition of alterations for each consecutive tumor. P2, P3 and P5 showed relatively short and neutral phylogenetic trees, suggesting weak evolutionary force. The short evolutionary tree of P5 could be related to a short time interval of only 1 year between the primary tumor and metastasis. Conversely, P2 and P3 have longer time intervals between the primary tumor and metastasis, ranging from 3 to 11 years. Comparing primary tumor samples before and after chemotherapy, samples were generally closely related to each other in the phylogenetic trees A great number of new alterations after neoadjuvant chemotherapy were only acquired in P1. The local recurrence of P1 also showed many unique alterations despite the relatively short time interval of 2 years between the primary tumor and the recurrence. This indicates a highly variable genome during tumor progression or the existence of many distinct subclones in this patient.

### Structural variants are the main alteration type and the frequency increases during progression

Absolute quantification of SNVs, indels and structural variants (translocations, deletions, duplications, insertions, inversions, SGLs and inferred structural variants) revealed that structural variants are the majority of alterations in osteosarcomas (figure 1B). In general, the most abundant structural variants types were single breakend structural variants, inversions and translocations. The frequency of insertions, SNVs and indels were least abundant. In all cases, the cumulative number of SNVs, indels and structural variants increased in later occurrences compared to the primary biopsy, with the exception of the first recurrence of P2 and the first metastasis of P4, where it slightly decreased. Proportionally, SNVs, indels and most SV types remained largely similar during tumor progression in P2, P3, P4, and P5 (figure 1B). Only P1 and P6 showed an increase of translocations and inversions, respectively, during progression. The consistent patterns of SV type proportions in four patients and the ongoing accumulation of INVs during progression in P6 might indicate the persistence of specific mutational mechanisms.

### Dynamic genome during progression characterized by the emergence of CNA and LOH

Analysis of copy number profiles showed that the majority of the osteosarcoma genomes were highly affected by CNAs and could show dynamic profiles during progression (figure 2A). Overall, P1, P2, P3, P4 and P6 harbored CNAs affecting short chromosome segments that showed small variations during progression. These short chromosome segments could suggest the presence of chromothripsis. Relatively stable copy number profiles were observed only for P5, where the majority of CNAs affected whole chromosome amplifications.

**Figure 2.**
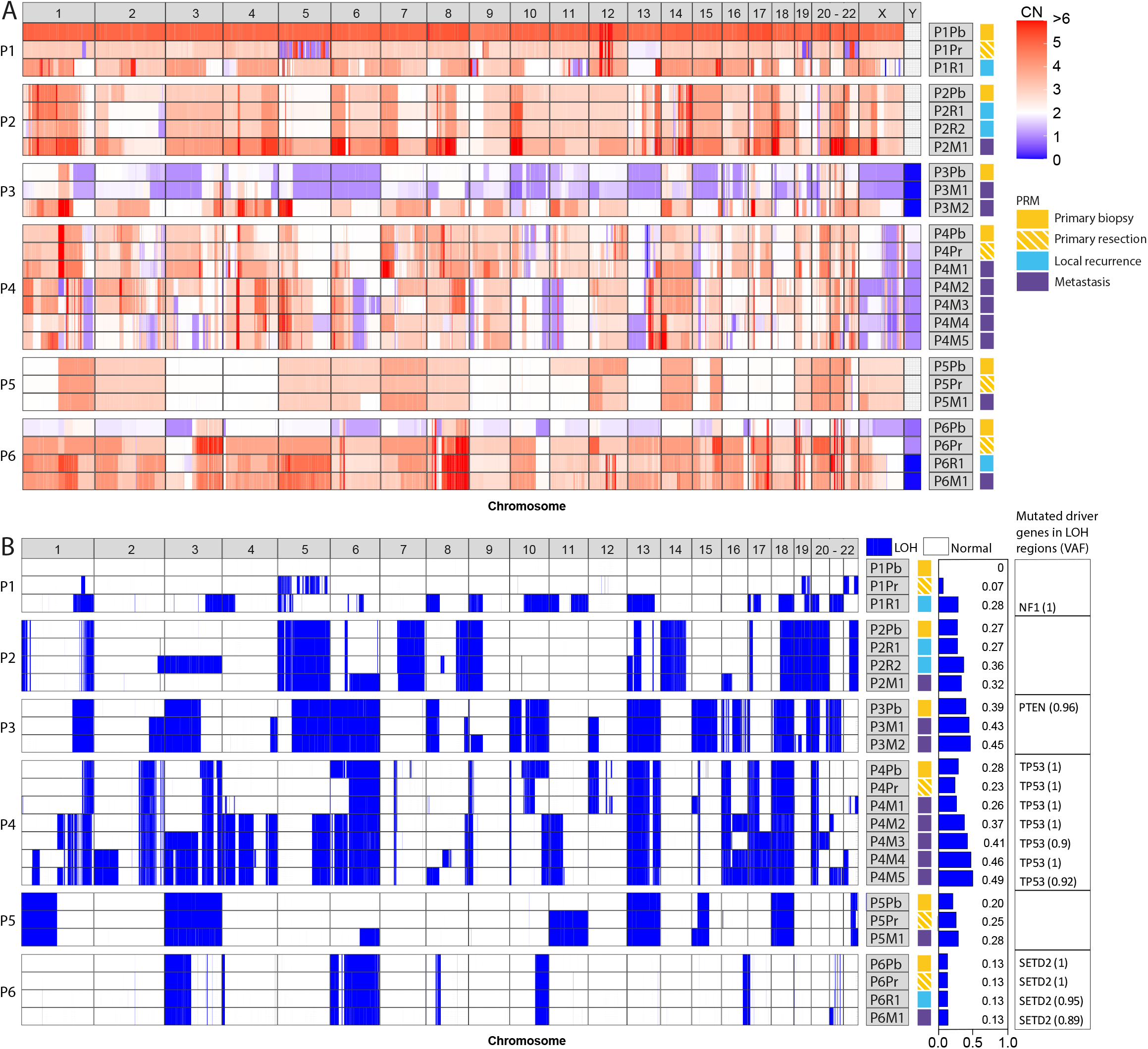
Copy number and LOH profiles during osteosarcoma progression. (A) Copy number profile displaying deleons (<2, blue), copy number neutral (2, white) and amplificaons (>2, red). (B) LOH profiles displaying regions of the genome affected by LOH (blue). The proporon of the enre genome affected by LOH and driver genes affected by LOH with the VAF of the mutaon are displayed next to the LOH profile. For both the copy number profile and LOH profile, paents are separated by white spaces and the tumors of each paent are ordered based on date of diagnosis. Chromosome numbers are noted on top of the copy number and LOH profiles.

Highlighting the interesting dynamic tumor genome of P1, each of the consecutive tumors show distinct copy number profiles. First of all, a high tumor ploidy of 5.2 was calculated for the primary biopsy, while the resection after chemotherapy and local recurrence had a lower ploidy of 2.9 and 3.25, respectively (supplementary table S1). The resection specimen showed shattered chromosomes 5 and 19 that resulted in an alternating pattern of amplifications and deletions, pointing towards the presence of chromothripsis. These alternating patterns are not retained in the local recurrence, in which different CNAs appear throughout the whole genome compared to the primary tumor samples. However, very specific amplifications in chromosome 12 are shared between all tumors of this patient. The genes *CDK4* and *MDM2* are located on those amplified regions and exhibited extremely high copy numbers (copy number of CDK4 and MDM2 respectively in: primary biopsy, 102 and 85; primary resection, 113 and 216; local recurrence, 28 and 93). P4 showed highly distinct copy number profiles when comparing the profiles of the primary tumor and first metastasis, as opposed to the profiles of the second metastasis through fifth metastasis, following the branched evolutionary pattern previously described.

Next to copy number profiles, loss of heterozygosity (LOH) can also be important for tumorigenesis. For example, both alleles can be inactivated in a two-step model where a mutation is acquired in one allele and the wild-type allele is lost due to LOH. Segments affected by LOH remained largely similar in osteosarcoma during progression in 5 out of 6 patients (figure 2B). Only P1 shows highly distinct LOH profiles for each tumor. As expected, regions with LOH are typically not restored as this is, in theory, not possible in clonally related cells. Regions where LOH profiles are restored in later occurrences would suggest the presence of multiple subclones. For example, the LOH of whole chromosome 3 in the second local recurrence of P2 is not detected in the following metastasis, suggesting that an earlier subclone seeded the metastasis. Potential driver genes affected by LOH included *NF1* in P1, *PTEN* in P3, *TP53* in P4 and *SETD2* in P6. However, only truncal alterations in *TP53* in P4 and *SETD2* in P6 were observed in all occurrences of the patient, indicating that these are more likely driver genes.

### WGD detected as early or late event

WGD was detected in all patients, although not all patients already showed WGD in the primary biopsy (supplementary table S1). The appearance of WGD in late occurrences was detected in the second metastasis of P3 and in the primary resection and later occurrences of P6. While the primary biopsy of P6 did not have WGD yet, chromosome arm 8q, which includes driver gene *MYC*, was already doubled. On top of the chromosome arm amplification, MYC was located in a region with extra focal amplifications, resulting in a *MYC* copy number of 9 and indicating a chromothripsis event. In later occurrences of this patient, the focal amplification of *MYC* increased (copy number of MYC: primary resection 57, local recurrence 77, metastasis 48). P2 already had WGD in the primary biopsy, but an elevated copy number and ploidy was observed in the metastasis.

### Chromothripsis events can accumulate or emerge in different subclones during progression

The variation in copy number profiles and the accumulation of many private structural variants during progression pointed towards the presence of chromothripsis events that could also impact different regions during osteosarcoma progression. Therefore, chromothripsis during osteosarcoma progression was further explored. Chromothripsis events were determined using ShatterSeek and visual inspection. Chromothripsis was observed in the primary biopsy of 5 out of 6 patients, (supplementary figure S1). Only P5 did not display any chromothripsis events. As described in previous research, osteosarcomas frequently harbor non-canonical chromothripsis events that are different from those found in other cancer types [6]. In typical chromothripsis events, segments oscillate between two copy number states, whereas in non-canonical chromothripsis events, the majority of the segments oscillate between three or more copy number states. Indeed, all detected were noncanonical events (percentage oscillating segments across all chromothripsis events: mean = 10.1%, SD = 10.4 %, range = 0.4% - 52.2%). In 3 out of the 6 osteosarcoma patients (P1, P2 and P4), various additional chromothripsis events were acquired and lost during progression. To give an example of accumulating chromothripsis, P1 showed a chromothripsis event in the primary biopsy in chromosome 12, leading to amplification of *MDM2* and *CDK4*, which was also retained in later occurrences (figure 3A). In the primary resection, chromothripsis events were already present that included interchromosomal rearrangements between chromosome 5, 12 and 19. Interestingly, only part of the intra- and interchromosomal rearrangements involving chromosome 5 and none of the rearrangements involving chromosome 19 were retained in the local recurrence. Instead, new chromothripsis events, involving chromosome 9, 17 and X, arose in the local recurrence. Another interesting chromothripsis pattern was observed in P4 (figure 3B). This patient displayed chromothripsis events affecting similar regions of multiple chromosomes in the primary biopsy, primary resection and first metastasis. However, the majority of these chromothripsis events were not observed anymore in metastasis 2 to 5. There, new chromothripsis events involving chromosome 1, 2, 5, 8, 13 and 15 were acquired. These distinct patterns also overlapped with the CNA patterns and branching of the phylogenetic tree based on structural variants and SNVs (figure 1A, 2A).

**Figure 3.**
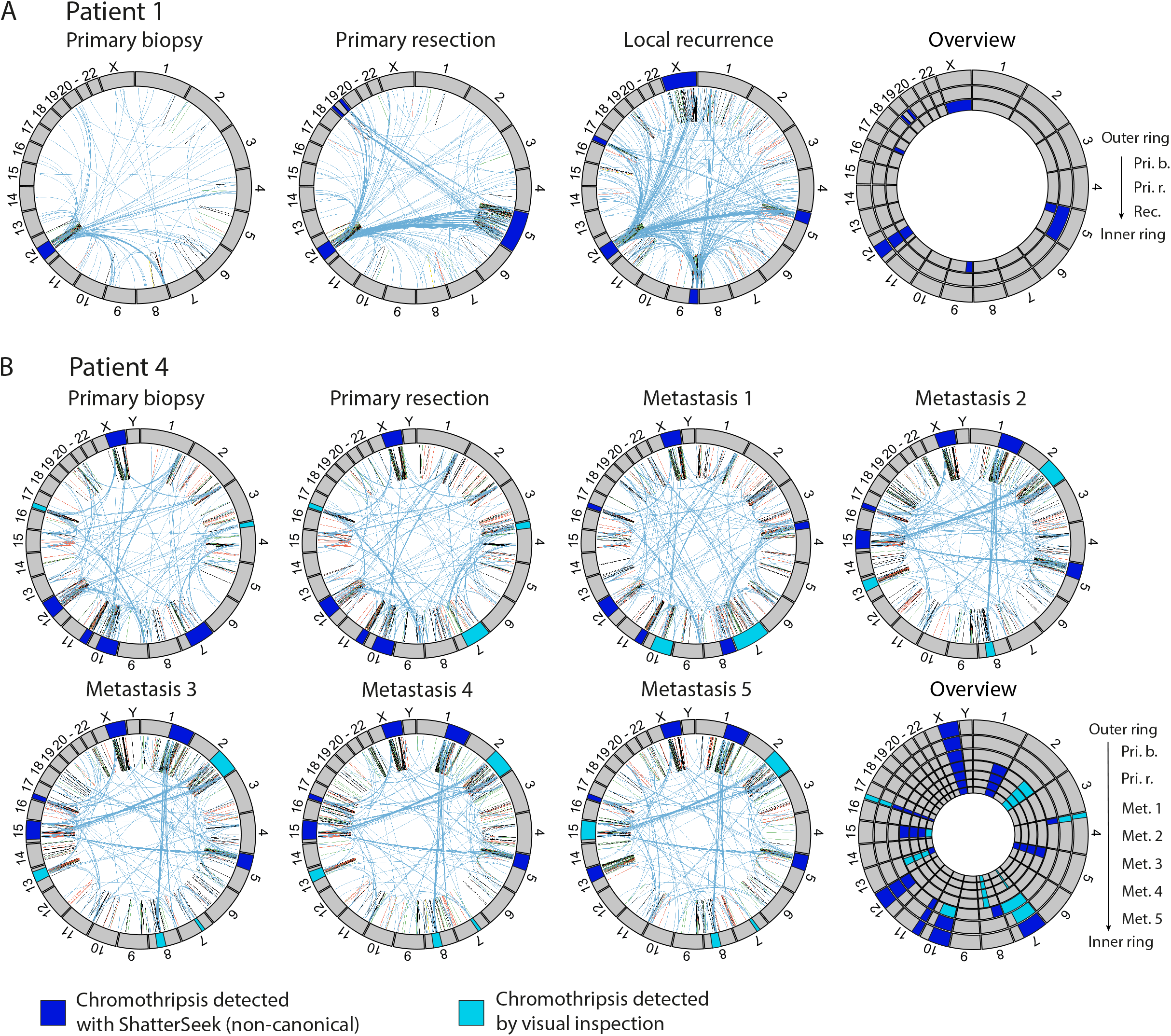
Presence of chromothripsis events in two osteosarcoma paents. Circos plots displaying accumulang chromothripsis events for paent 1 (A) and paent 4 (B). Chromosome numbers are annotated on the outer ring of the circos plot. Regions of the genome affected by chromothripsis are annotated by dark blue (calls with ShaerSeek) and light blue (calls by visual inspecon) bars in the chromosome annotaon ring. The inner circle displays the SVs and the SV type found in each tumor: translocaons (blue), deleons (red), inserons (yellow), duplicaons (green) and inversions (black). The overview circos plot summarizes the chromothripsis events of all occurrences of the paent.

### Colocalization of potential driver genes and chromothripsis events

To further investigate the impact of chromothripsis, we looked whether chromothripsis regions co-localize with the potential driver genes. In P1, a *CDK4* and *MDM2* amplification was found which colocalized with a chromothripsis event that was found in all occurrences of this patient (figure 4A). Alterations in *TP53* were found in P2, P3 and P4. Interestingly, chromothripsis in chromosome 17 was not detected in P2 and P3, despite the presence of structural variants in *TP53* leading to a translocation in P2 and a deletion in P3. In P4, *TP53* localized in the middle of the chromothripsis event (figure 4B). Furthermore, a deletion-like event spanning over *TP53* was observed, explaining the LOH. In P6, chromothripsis events observed in chromosome 8 colocalized with *MYC* (figure 4C). Additionally, two chromothripsis events occurred in chromosome 17, although these events did not colocalize with *TP53*.

**Figure 4.**
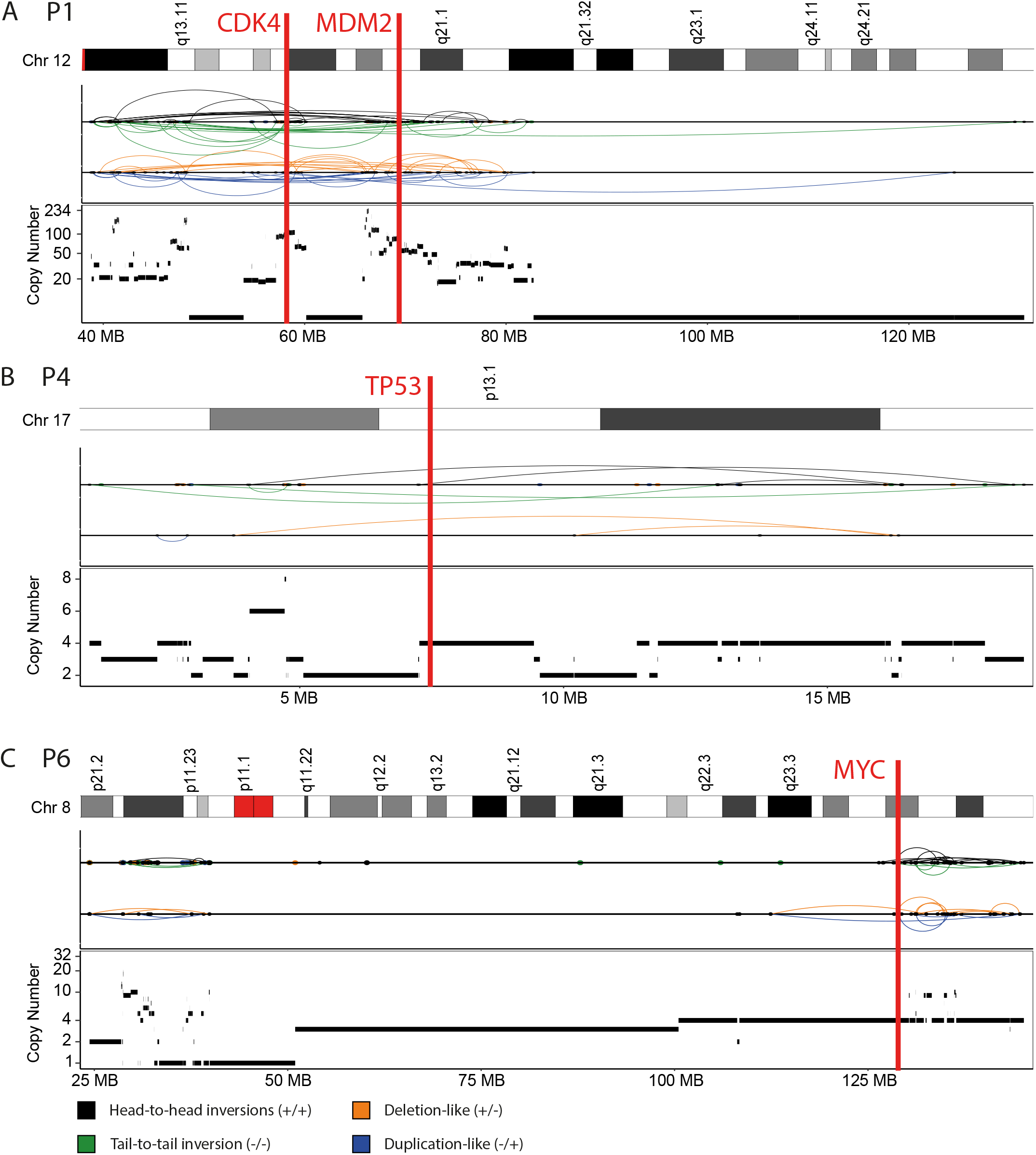
Colocalizaon of driver genes and chromothripsis events. ShaerSeek visualizaon of chromothripsis events in (A) P1 chromosome 12, (B) P4 chromosome 17 and (C) P6 chromosome 8. Locaon of selected driver genes are annotated with a red bar.

### Therapeutically actionable targets

Pathogenic mutations can serve as targets for personalized treatment in specific cancer types and anticancer drugs against pathogenic mutations have also shown efficacy outside their approved label [16]. Therefore, the potential driver genes were screened for therapeutically actionable targets using the OncoKB database. Only five actionable targets could be identified, including the *MDM2* amplification in all tumors of P1, an *ATR* mutation in the primary biopsy of P1, a *NF1* mutation in the local recurrence of P1, a *PTEN* mutation in the primary biopsy of P3 and a *PTEN* mutation in the primary resection of P4 (supplementary table S1). The *MDM2* amplification and *NF1* were found to be the only stable alterations as the *MDM2* amplification was retained throughout all tumors of P1 and *NF1* had a variant allele frequency of 1 in the local recurrence of P1. Conversely, the *ATR* and two *PTEN* mutations were found to be unstable as they were exclusively observed in the primary tumor, making them ineligible as targets for therapy. Thus, only two out of six patients WGS provided potential targets for therapy.

## Discussion

The extreme genomic heterogeneity between and within osteosarcoma patients underlines the difficulty to understand osteosarcoma initiation and progression. In our study, we performed a detailed characterization of the osteosarcoma genome from 6 patients during progression using WGS. In-depth analysis of SNVs, structural variants, CNAs and chromothripsis showed that the majority of osteosarcoma genomes are highly variable and dynamic during disease progression.

Longitudinal analysis of genomic alterations in osteosarcoma genome revealed truncal mutations in genes that could initiate and drive tumor development. As expected, *TP53* was the most frequently mutated truncal driver gene among the patients, although it was affected in different ways in each patient, including a gene fusion, deletion and a SNV combined with LOH. Interestingly, a recent study showed that structural variants in *TP53* in osteosarcoma commonly result in loss of coding parts while preserving and relocating the gene’s promoter region [17]. However, this type of *TP53* alteration was not observed here. Other truncal mutations in potential driver genes involved the simultaneous high level amplification of *CDK4* and *MDM2* that co-localized with chromothripsis. Notably, these amplifications were previously also linked to chromothripsis in liposarcoma [18]. Mutations arising later during disease progression in potential driver genes identified in this study, such as *TERT* and *PDGFRA*, could have contributed to disease progression. For example, *TERT* promoter mutations have frequently been found in chondrosarcomas, the second most common primary bone sarcoma after osteosarcoma. The *TERT* mutations occur more often in high grade chondrosarcoma and dedifferentiated chondrosarcoma compared to low grade chondrosarcoma and is associated with a poor survival, indicating that *TERT* could play an important role in tumor progression [19, 20]. Furthermore, studies have shown that overexpression of *TERT* is associated with decreased progression free survival and promotes chemoresistance in osteosarcoma [21, 22]. The PDGF/PDGFR pathway is thought to have an important role in the development of osteosarcoma metastases [23]. High expression of *PDGF-AA* is associated with short disease-free survival in osteosarcoma compared to low *PDGF-AA* expression and a similar trend was observed for *PDGFRA* expression [24]. Additionally, the simultaneous amplification of *PDGFRA, KIT* and *KDR*, clustered on 4q12, is found to be a significant event across different cancer types and is particularly enriched in osteosarcoma [25]. Moreover, multiple studies using multi-kinase inhibitors against PDGFRA and KDR activity have shown clinical activity in advanced osteosarcoma [26-28]. Comparing the genome of chemotherapy naïve tumors with tumors after chemotherapy treatment, we found that these tumors were closely related in the phylogenetic trees in three out of four patients that had genomic data of both tumors. While previous research found that mutations in *NF1* and *KIT* could be potentially acquired after chemotherapy [9], we did not find these mutations after chemotherapy, although they were observed in later occurrences of two patients.

Compared to SNVs and indels, the large number of structural variants had a substantial contribution to the shape of the evolutionary trees. Previously described evolutionary trees that are based only on SNVs and indels already showed intra- and inter-patient heterogeneity. However, these could not capture evolutionary patterns driven by structural variants [8, 10]. By taking into account structural variants, highly distinct phylogenetic tree patterns were observed, which further emphasized the inter- and intra-patient heterogeneity in osteosarcoma. Moreover, intra-patient heterogeneity was also observed in copy number profiles. Variable copy number profiles were detected in five out of six patients during disease progression. In contrast, a previous study looking at CNAs in single cells during osteosarcoma progression only identified little intra-patient heterogeneity, although this was observed in a PDX model, in which only a small piece of tumor was clonally grown with a limited timeframe [29].

The accumulation of many private structural variants in metastases suggested that novel chromothripsis events can arise in later occurrences as well. Our longitudinal chromothripsis analysis confirmed this. Additionally, we noticed that all chromothripsis events in osteosarcomas harbor different characteristics compared to canonical chromothripsis events seen in most other cancers, as was previously described [6]. Interestingly, few disappearances of chromothripsis events and LOH regions were observed as well, suggesting the presence of highly distinct subclones. While this study only allowed the sequencing of single-region samples due to the scarcity of tumor material, a subclonal reconstruction with a multi-region sampling design could further elucidate the existence of subclonal chromothripsis [30].

The highly variable osteosarcoma genome poses a great challenge in the treatment of this disease. Osteosarcomas generally have only a few actionable targets which can be unstable similar to the rest of the osteosarcoma genome and are therefore not optimal as targets for therapy. While targeted therapy is successful in other cancer types [16], targeted therapy towards a single molecular variant might not have the same potential in osteosarcoma. Therefore future research could focus on treatment options from different perspectives, such as harnessing the tumor microenvironment.

To conclude, our comprehensive longitudinal genomic analysis of osteosarcoma revealed the ongoing gain and loss of large genomic events during osteosarcoma progression. These findings provide more insight into the extreme degree of osteosarcoma inter-and intra-patient heterogeneity and underscores the complexity of osteosarcoma development and treatment.

## Data Availability

All data produced in the present study are available upon reasonable request to the authors

## Author contributions

**D.M. Meijer**: Conceptualization, methodology, project administration, data curation, software, formal analysis, validation, investigation, visualization, writing-original draft, writing-review and editing. **D. Ruano**: Investigation, writing-review and editing. **I.H. Briaire-de Bruijn**: Investigation, writing-review and editing. **P.M. Wijers-Koster**: Investigation, writing-review and editing. **M.A.J. van de Sande**: Resources, writing-review and editing. **H. Gelderblom**: Resources, writing-review and editing. **A.M. Cleton-Jansen**: Investigation, writing-review and editing. **N.F.C.C. de Miranda**: Conceptualization, methodology, supervision, writing-review and editing. **M.L. Kuijjer**: Conceptualization, methodology, supervision, writing-original draft, writing-review and editing. **J.V.M.G. Bovée**: Conceptualization, methodology, supervision, writing-original draft, writing-review and editing.

## Ethics approval and consent to participate

Approval of institutional review board (Medisch-Ethische Toetsingscommissie Leiden Den Haag Delft) was obtained for the use of tissue samples from the LUMC bone and soft tissue tumor biobank (Leiden, the Netherlands) (B20.067). Written informed consent from patients was obtained. All specimens were pseudoanonymized and handled according to the ethical guidelines described in ‘Code for Proper Secondary Use of Human Tissue in The Netherlands’ of the Dutch Federation of Medical Scientific Societies.

## Competing interests

The authors declare no conflict of interest.

## Funding information

This work was financially supported by the intramural Leiden Center for Computational Oncology strategic fund. M.L.K.is funded by the Norwegian Research Council, Helse Sør-Øst and the University of Oslo through the Centre for Molecular Medicine Norway (grant no. 187615), the Norwegian Research Council (grant no. 313932), and the Norwegian Cancer Society (grant no. 214871 and 273592). N.F.C.C.d.M. is funded by the European Research Council (ERC) under the European Union’s Horizon 2020 Research and Innovation Program (grant agreement no. 852832) and by the VIDI ZonMW (project number: 09150172110092).

## Notes

### Competing Interest Statement

The authors have declared no competing interest.

### Author Declarations

Approval of institutional review board (Medisch-Ethische Toetsingscommissie Leiden Den Haag Delft) was obtained for the use of tissue samples from the LUMC bone and soft tissue tumor biobank (Leiden, the Netherlands) (B20.067). Written informed consent from patients was obtained. All specimens were pseudoanonymized and handled according to the ethical guidelines described in Code for Proper Secondary Use of Human Tissue in The Netherlands of the Dutch Federation of Medical Scientific Societies.

