## Supplementary_figure_S1 for "The variable genomic landscape during osteosarcoma progression: insights from a longitudinal WGS analysis"

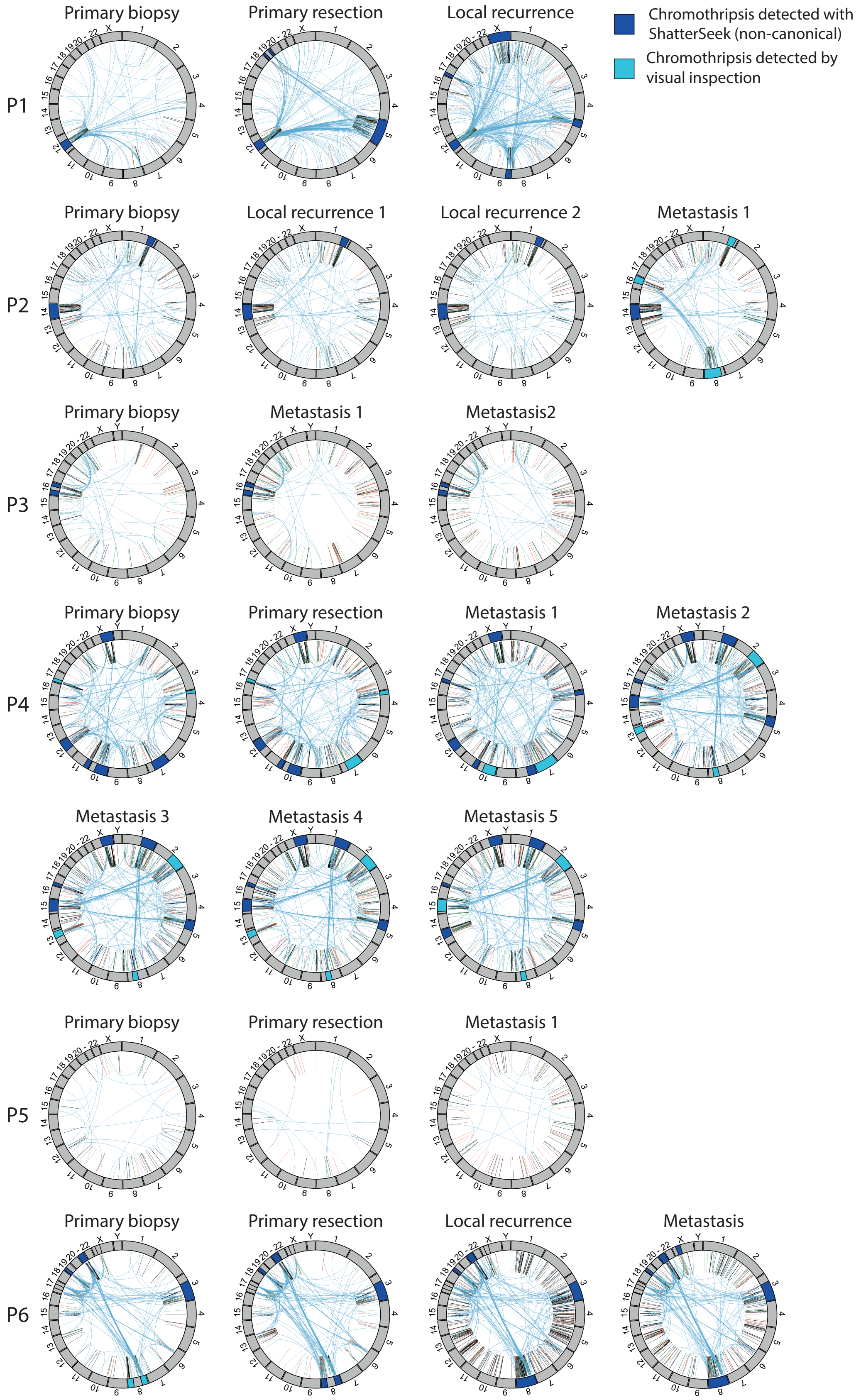


**Supplementary figure 1.** Circos plots displaying the chromothripsis events in all patients. Chromosome numbers are annotated on the outer ring of the circos plot. Regions of the genome affected by chromothripsis are annotated by white (calls with ShatterSeek) and black (calls by visual inspection) bars in the chromosome annotation ring. The inner circle displays the SVs and the SV type found in each tumor: translocations (blue), deletions (red), insertions (yellow), duplications (green) and inversions (black).
